# "Spatial Disparities and Multilevel Determinants of Diarrhea Among Under-Five Children in Mozambique: Evidence from the 2022/2023 Demographic and Health Survey"

**DOI:** 10.1101/2025.08.01.25332728

**Authors:** Thomas Kidanemariam Yewodiaw, Mihret Getnet, Mequanent Dessie Bitewa, Hiwot Tezera Endale

## Abstract

**Background:** Diarrhea remains a major health challenge in Mozambique, with its burden varying across regions. An updated, detailed analysis using the 2022/2023 DHS data is essential for informed public health planning and prioritize resource allocation. This study explores the spatial distribution and key determinants of diarrhea among children under five to design targeted interventions.

**Methods:** This study used data from 9,799 under-five children in the 2022/2023 Mozambique DHS to assess diarrhea prevalence and determinants. Weighted analysis, SaTScan, hotspot mapping, and multilevel logistic regression were used to assess individual and community factors influencing diarrhea among under-five children. Significant predictors were identified using AORs (95% CI, *p* ≤ 0.05). ICC, MOR, and PCV assessed model variation, and spatial patterns were mapped using ArcGIS.

**Results:** The weighted prevalence of diarrhea among under-five children in Mozambique was 8.8% (95% CI: 7.8–9.6), with the highest rates in Niassa (14.5%), Cabo Delgado (14.0%), and Maputo City (13.4%), and the lowest in Maputo Province (5.1%), Zambézia (5.5%), and Manica (5.8%). Multilevel analysis revealed children aged 12–23 months had significantly higher odds of experiencing diarrhea (AOR = 1.36, 95% CI: 1.11–1.66), while those aged 24–59 months (AOR = 0.49, 95% CI: 0.41–0.59), compared to infants aged 0–11 months, Maternal education (AOR = 0.77, 95% CI: 0.63–0.96), Rural residence (AOR = 0.69, 95% CI: 0.52–0.91) were protective, and children residing in regions such as Nampula (AOR = 0.42), Zambézia (AOR = 0.35), Manica (AOR = 0.30), Sofala (AOR = 0.51), and Maputo Province (AOR = 0.26) showed significantly lower odds compared to Niassa. Health-seeking behavior showed a strong positive association with diarrhea occurrence (AOR = 4.85, 95% CI: 4.10–5.74).

**Conclusions:** Diarrhea among under-five children in Mozambique shows marked regional variation, with higher rates in Niassa, Cabo Delgado, and Maputo City, and lower rates in Maputo Province, Zambézia, and Manica. Risk was linked to child age, maternal education, and region, while rural residence and certain regions were protective. Targeted, region, and age-specific interventions are needed.

## Introduction

Diarrheal Disease is the third leading cause of death in children 1-59 months of age particularly in low income and rural settings. But, it is both preventable and treatable (1). According to the most recent WHO and UNICEF data from 2024-2025, Each year approximately 444,000 children under five years old die globally(1, 2) and 2021 estimates suggests around 340,000 to 390,000 child deaths from diarrhea, reflecting some variations depending on data sources and methods(1, 3). Nearly 1.7 billion cases of childhood diarrhea occur every year worldwide(1).

The prevalence of diarrhea among children under five years old in East Africa and Sub-Saharan countries was 14.28% and 15.3% respectively(4, 5). The 2011 Mozambique demographic and health survey revealed a 11% prevalence of childhood diarrhea, with higher rates in teta and Zambia, but lower rates in Gaza and Cabo Delgado(6). UNICEF (2021) highlights diarrhea as a major cause of under-five mortality in Mozambique, exacerbated by inadequate access to clean water, poor sanitation, and limited healthcare services(7). In Mozambique, diarrhea continues to be a moderate but persistent public health concern, especially among children under five(8). Previous surveys have revealed geographical inequalities, with higher frequency in northern regions, frequently connected to insufficient water and sanitation, low maternal education, and limited healthcare access(9, 10). The inequalities underscore the need to analyze both individual-level factors like child age and maternal employment and community-level determinants like region and rural/urban setting (11).

Childhood diarrhea risk factors include child age, breastfeeding status, vaccination status, nutritional status, household size, caregiving behavior, maternal age, education level, water source, sanitation, socioeconomic status, and vaccination coverage (12-16).

DHS data-driven spatial epidemiological analyses help identify high-risk geographic clusters for childhood diarrhea, enabling policymakers to allocate resources and implement intervention strategies in high-burden areas(17). Mozambique, a southeastern African country, is significantly burdened by childhood diarrhea, with high prevalence in children under five, varying across provinces. Regarding to SDG goal three the Mozambique planned to reduce under five children mortality by 25/1000 (18, 19). The 2022/2023 Mozambique Demographic and Health Survey (DHS) offers recent, nationally representative data to reassess diarrhea prevalence and patterns across the region(8, 20). The analysis of recent public health initiatives and changes in infrastructure and health services is crucial to identify changes in disease distribution and risk factors(18). Understanding the current burden and predicator of diarrhea is vital to inform evidence-based, age- and region-specific interventions(5). The study aims to evaluate diarrhea prevalence, spatial distribution, and associated factors among under-five children in Mozambique using multilevel logistic regression and spatial analysis (e.g., SaTScan and hotspot mapping)(21). The findings will aid national efforts to decrease childhood morbidity and contribute to achieving Sustainable Development Goals related to child health and water and sanitation (SDG 6) (22).

## Methods and materials

### Study setting

Mozambique, located in south-east Africa at 18° 15’ South and 35° 00’ East, has an estimated 34 million people, the bulk of whom live in rural areas(23-25). The country borders the Indian Ocean, Tanzania, Malawi, Zambia, Zimbabwe, and Eswatini and South Africa. Mozambique is divided into 10 provinces and one city province (Maputo City)(26).

### Data source and sampling procedure

The analysis utilizes Mozambique 2022/2023 Demographic and Health Survey data (DHS) sourced from the MEASURE DHS public repository. The study utilized the latest data set from the DHS, a worldwide survey conducted every five years in low- and middle-income nations. The data was gathered from a national representative sample of approximately households from all 11 regions of Mozambique. The 2022/2023 Mozambique demographic and health survey utilized stratified two-stage cluster sampling to provide representative results at the national level for urban and rural areas. The 619 targeted clusters were selected using a probability-proportional-to-size strategy for urban and rural areas, followed by systematic random sampling for equal probability. The second stage involves updating household listings and maps in all selected clusters to create a list of households for each cluster. The household sample was drawn from the list. From the list, a predetermined number of 30 households per cluster were chosen at random to participate in interviews. A study interviewed households 9,799 in 619 clusters, including 4,767 male and 5,032 female aged 0-59 months, with rural and urban. The study looked at 9,799 children aged 0 to 59 months, concentrating on the past two weeks of diarrhea history before to the survey.

### Study population

The study involved children 0-59 months aged in selected areas, with mothers interviewed for the survey, and mothers with multiple children were asked about their most recent child.

### Inclusion and exclusion criteria

The study included children aged 0-59 months in selected enumeration areas (EAs), while those aged 0-59 months not assessed for recent diarrhea or with missing outcome variables were excluded.

### Study variables

#### Outcome of variable

The study analyzed diarrheal illness in under-five children aged 0-59 months, coding it as "Yes=1" for recent diarrhea and "No=0" for no recent diarrhea.

#### Independent variables

Those factors were reviewed from different literatures, include child age, breastfeeding status, vaccination status, nutritional status, household size, caregiving behavior, maternal age, education level, residence, region, water source, sanitation, and socioeconomic status(12-16).

### Operational Definitions

#### Diarrhea

A child is considered to have diarrhea if the mother or caregiver reports that the kid passed three or more loose or watery stools in a 24-hour period within the two weeks preceding the survey(20).

#### Media Access

A respondent is considered to have media access if they report accessing any one of these media sources at least occasionally (newspapers or magazines, radio, or television)(20).

#### Types of toilets

Toilets such as flush to elsewhere, pit latrines without slabs, bucket, hanging, or other types were classified as **unimproved**. In contrast, flush toilets connected to sewers, septic tanks, pit latrines, ventilated or slab pit latrines, and composting toilets were considered **improved sanitation facilities(20, 27, 28)**.

#### Source of Water

drinking water sources are classified as **improved** if they include piped water, public taps, tube wells, protected wells or springs, rainwater, and bottled water. Sources like unprotected wells or springs, tanker trucks, surface water, and bottled water used alone are considered **unimproved(20, 27).**

**Wealth Index** is a combined measure of a household’s overall living conditions, based on ownership of assets (TV, bicycle), housing quality (floor material, water source, sanitation facilities), and access to services. Using principal components analysis, DHS assigns weights to these factors and classifies households into five groups, ranging from poorest to richest(20, 29).

### Data management and analysis

The study utilized data from various sources, including median, table, and percent, to analyze key characteristics and estimate response rates and frequencies due to non-proportional sample allocation. The weighting procedure can be explained in more detail on the DHS portal at https://dhsprogram.com/data/Guide-to-DHS-Statistics/Analyzing_DHS_Data.htm. Figures were created using ArcGIS version 10.7 software, using Mozambique regional shape file for hotspot area, kriging interpolation, and SatScan windows, available at https://data.humdata.org/dataset/cod-ab-moz

### Spatial analysis

The spatial analysis of under-five diarrhea in Mozambique was conducted using ArcGIS 10.7 and Sat Scan version 9.6, using Global Moran’s I statistics measure to evaluate its distribution (dispersed, clustered, or randomly). Moran’s I value is a spatial statistic used to measure spatial autocorrelation by generating a single value from -1 to 1. Moran’s I value indicates a clustered pattern of under-five diarrhea, a negative value indicates a dispersed pattern, and a close to zero value indicates a random distribution(30, 31).

**Getis-Ord Gi* statistics hotspot analysis(32)** was used hot spot analysis to identify the hotspot and cold spots in the cluster, indicating a higher or low proportion of under-five diarrhea illness. Hotspot and cold spot cluster are indicated high and low proportions of diarrhea among under five children respectively. The spatial interpolation technique was employed to predict diarrheal illnesses among under five children in unsampled areas based on sampled EA measurements. This study utilized the ordinary Kriging spatial interpolation method, which has the smallest root mean square error value and residuals, to predict diarrheal illnesses in unobserved areas(33). The study utilized spatial scan statistical analysis (SaTScan) with the Bernoulli distribution to identify significant spatial clusters of childhood diarrheal illnesses using Kulldorf’s SaTScan V.9.6 software(34).

### Characteristics of study participants in Mozambique

#### Multilevel Analysis

The multilevel multivariable logistic regression model was used to analyze the association between individual and community-level factors, with fixed effect estimates and a 95% confidence interval. Multilevel analysis is appropriate for nested data, such as DHS. Individual-level factors include child age, sex, mother’s education, mother employments, mother age, household wealth, Household size, and nutritional status are associated risk of diarrhea among children. However, Mothers’ individual characteristics, like as income and education, may be influenced by community-level factors such as place, media exposure, poverty, and region. Individual-level characteristics of children in DHS data tend to be more connected within clusters than between other clusters. The similarity of individual characteristic scores within a cluster violates conventional regression’s independence assumptions. Multilevel analysis can overcome the lack of independence of observations in nested data analysis. A two-level binary logistic regression (individual and community level) was used to determine risk variables for diarrhea.

A total of four models were fitted. The null model, commonly known as the random intercept model, was used to assess cluster variability in diarrhea. Model fit was determined using fitness requirements such as the Likelihood Ratio test (LLR), deviance, Akaike information criterion (AIC), and Deviance Information Criterion (DIC). The model with the lowest fitness parameters was chosen as the optimal fit.

The study evaluated cluster variability using the intra-class coefficient (ICC)(35) and median odds ratio and Proportional Change in Variance (PCV). The ICC measures the percentage variation in community-level variables, while PCV measures the proportional change in community-level variance between null and succeeding models(36). The Median Odds Ratio (MOR) is a statistical measure that measures the area-level variance of the odds ratio scale. It is calculated by comparing the median value of the odds ratio between high and low risk areas. In the absence of area-level variation, the MOR is equal to 1. Adjusted odds ratio with 95% confidence interval (CI) and p-value < 0.05 were used to declare statistical significance.

### Ethical considerations

This study utilized secondary data from the 2022/2023 Mozambique Demographic and Health Survey (DHS), which is publicly available and fully anonymized. The original DHS data collection was approved by the Institutional Review Board of ICF and the Mozambique National Committee for Bioethics in Health (CNBS), and informed consent was obtained from all participants at the time of the survey.

Access to the dataset for this analysis was granted by the DHS Program on February 2, 2024, under a previous project request. As this is a secondary analysis of de-identified, publicly available data, no additional ethical approval was required for the current study.

Although public access to DHS datasets is currently paused, the data used in this study were obtained through an earlier approved request. Interested researchers can access the data upon reasonable request and approval from the DHS Program via https://dhsprogram.com.

## Results

### Characteristics of the study population

The majority of under-five children were aged 24–59 months, accounting for 60% (5,881) of the sample. Females slightly outnumbered males at 51.4% (5,032) compared to 48.6% (4,767). Most mothers had primary education 49.1% (4,810), while 30.1% (2,950) had no formal education. The dominant maternal age group was 20–34 years 69.5% (6,812). A large proportion of mothers were not working 73.2% (7,172). The sample was mostly rural 71.3% (6,991). In terms of wealth, 48.1% (4,709) of children were from low-income households. Nearly half lived in small-sized households 48.8% (4,783). Nutritional status was largely normal 94% (9,214), while underweight cases were 6% (585). Only 25.3% (2,483) of children were fully vaccinated. Most children lived in communities with low underweight prevalence 98.7% (9,670), and a majority lived in medium 37.5% (3,673) or high 35.2% (3,452) education-level communities (Table 1).

**Table 1:**
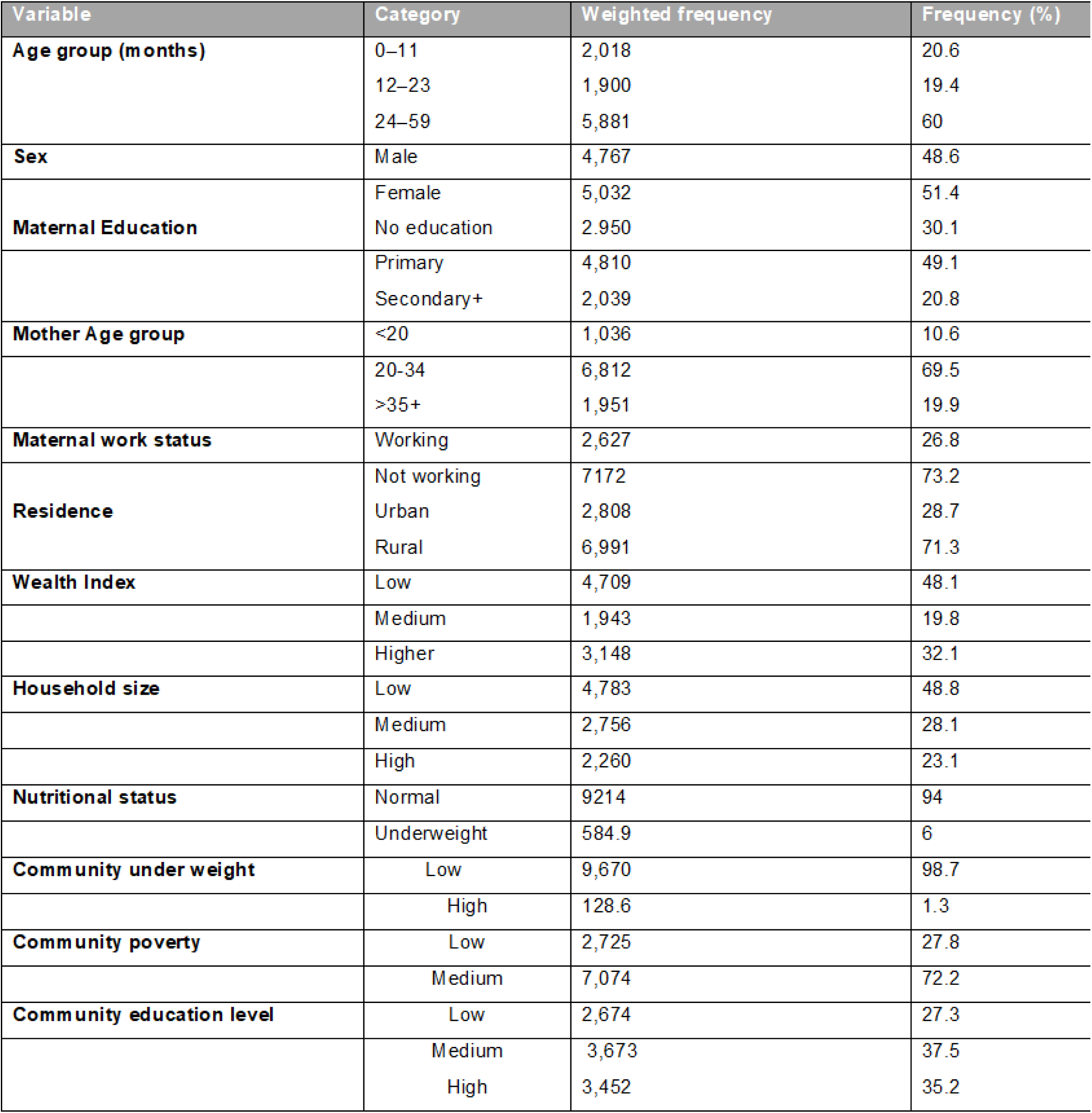
Characteristics of the study population.

### National Prevalence of Diarrhea among Under-Five Children

The overall prevalence of diarrhea among under-five children in the 2022/2023 Mozambique DHS KR dataset was 8.8% (95% CI: 7.8% to 9.6%) Weighted case= 817 children) during the two weeks preceding the survey. Diarrhea was more common among children aged 12-23 months, with a prevalence of 15.1% (95% CI: 13% to17.5%), and was more frequently reported among urban (11.7%). The burden was higher among children of mothers with secondary education and above (10.9%) (Table 2).

**Table 2:** Weighted Prevalence of Diarrhea Among Under-Five Children by Selected Characteristics (Mozambique DHS 2022/2023.

| *Variable* | *Category* | *Prevalence (%)* | *95% CI* | *Weighted N* |
| --- | --- | --- | --- | --- |
| **Child Age (months)** | *0–11* | *10.4* | *(8.7 – 12.3)* | *202* |
|  | *12–23* | *15.1* | *(13 – 17.5)* | *272.9* |
|  | *24–59* | *6.1* | *(5.3 – 7)* | *342.2* |
| **Sex** | *Male* | *8.5* | *(7.4 – 9.7)* | *385.4* |
|  | *Female* | *8.9* | *(7.9 – 10.1)* | *431.7* |
| **Place of Residence** | *Urban* | *11.7* | *(10.2 – 13.4)* | *315* |
|  | *Rural* | *7.5* | *(6.5 – 8.6)* | *502.1* |
| **Maternal working status** | *Working* | *10.9* | *(9.4-12.6)* | *277.9* |
|  | *Not working* | *7.9* | *(6.9-8.9)* | *539.2* |
| **Community Poverty** | *Low* | *11* | *(9.6 – 12.4)* | *288.8* |
|  | *Medium* | *7.8* | *(6.8 – 9)* | *528.3* |

### Regional prevalence of Diarrhea in Under-Five Children

*The 2022/2023 DHS data reveals the prevalence of diarrhea among under-five children in Mozambique shows noticeable regional variation. The two provinces with the **highest diarrhea prevalence** are **Niassa** (14.6%) and **Cabo Delgado** (14.1%), In contrast, the **lowest prevalence** is observed in **Maputo Province** (5.2%) and **Zambézia** (5.5%) (Table 3)*.

**Table 3:**
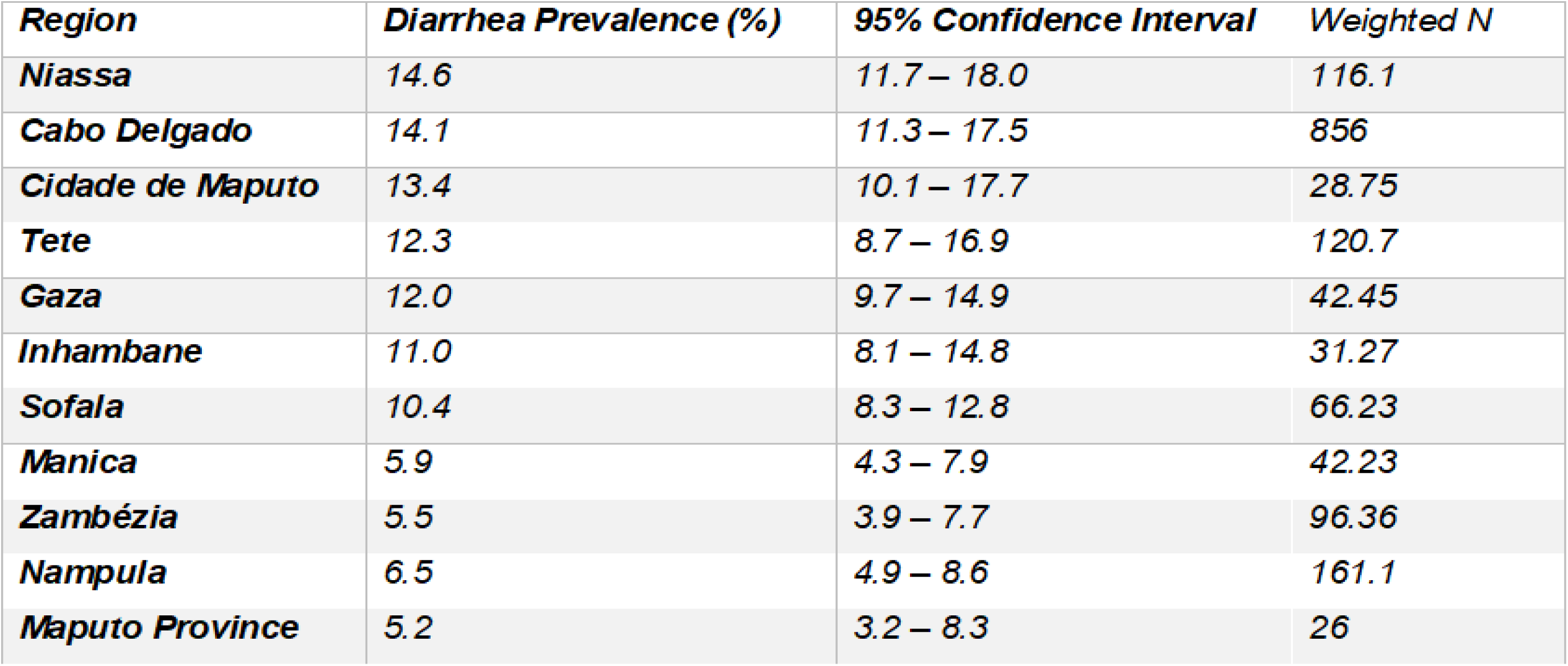
Prevalence of Diarrhea Among Under-Five Children by Region – Mozambique DHS 2022/2023.

| **Region** | **Diarrhea Prevalence (%)** | **95% Confidence Interval** | **Weighted N** |
| --- | --- | --- | --- |
| **Niassa** | 14.6 | 11.7 – 18.0 | 116.1 |
| **Cabo Delgado** | 14.1 | 11.3 – 17.5 | 856 |
| **Cidade de Maputo** | 13.4 | 10.1 – 17.7 | 28.75 |
| **Tete** | 12.3 | 8.7 – 16.9 | 120.7 |
| **Gaza** | 12.0 | 9.7 – 14.9 | 42.45 |
| **Inhambane** | 11.0 | 8.1 – 14.8 | 31.27 |
| **Sofala** | 10.4 | 8.3 – 12.8 | 66.23 |
| **Manica** | 5.9 | 4.3 – 7.9 | 42.23 |
| **Zambézia** | 5.5 | 3.9 – 7.7 | 96.36 |
| **Nampula** | 6.5 | 4.9 – 8.6 | 161.1 |
| **Maputo Province** | 5.2 | 3.2 – 8.3 | 26 |

### Spatial analysis

#### Spatial autocorrelation

The geographical autocorrelation analysis indicated regional variation in diarrhea among under-five children in Mozambique. The Global Moran’s I was 0.05, with a Z-score of 1.74 and a p-value of 0.08. While these values suggest a weak positive spatial autocorrelation, the result is not statistically significant at the 5% level. Therefore, although there may be a slight tendency toward geographic clustering, we cannot confidently conclude that the spatial distribution of diarrhea is non-random. Further localized spatial analysis is recommended to explore potential clustering at sub-regional levels

**Figure 1:**
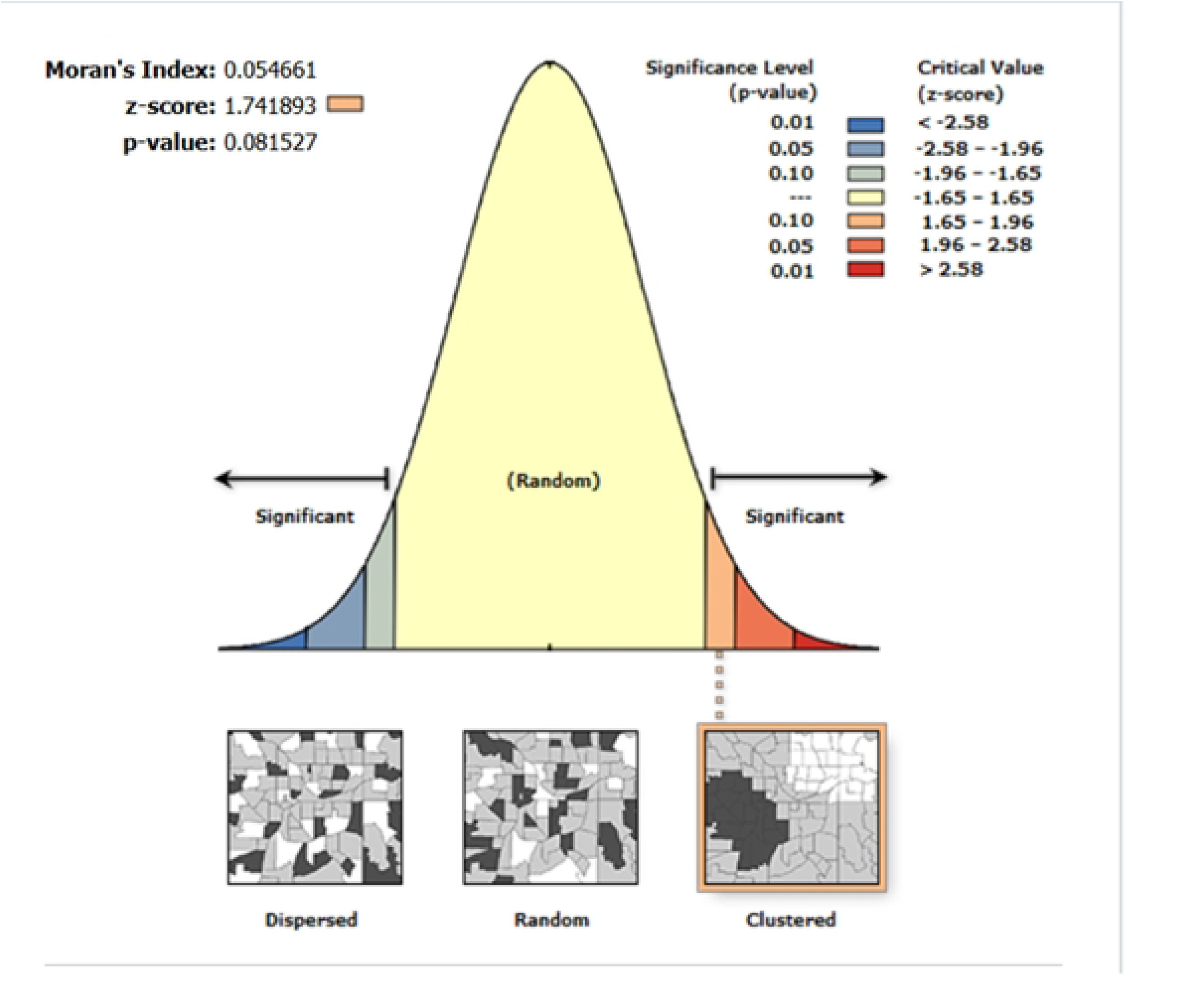
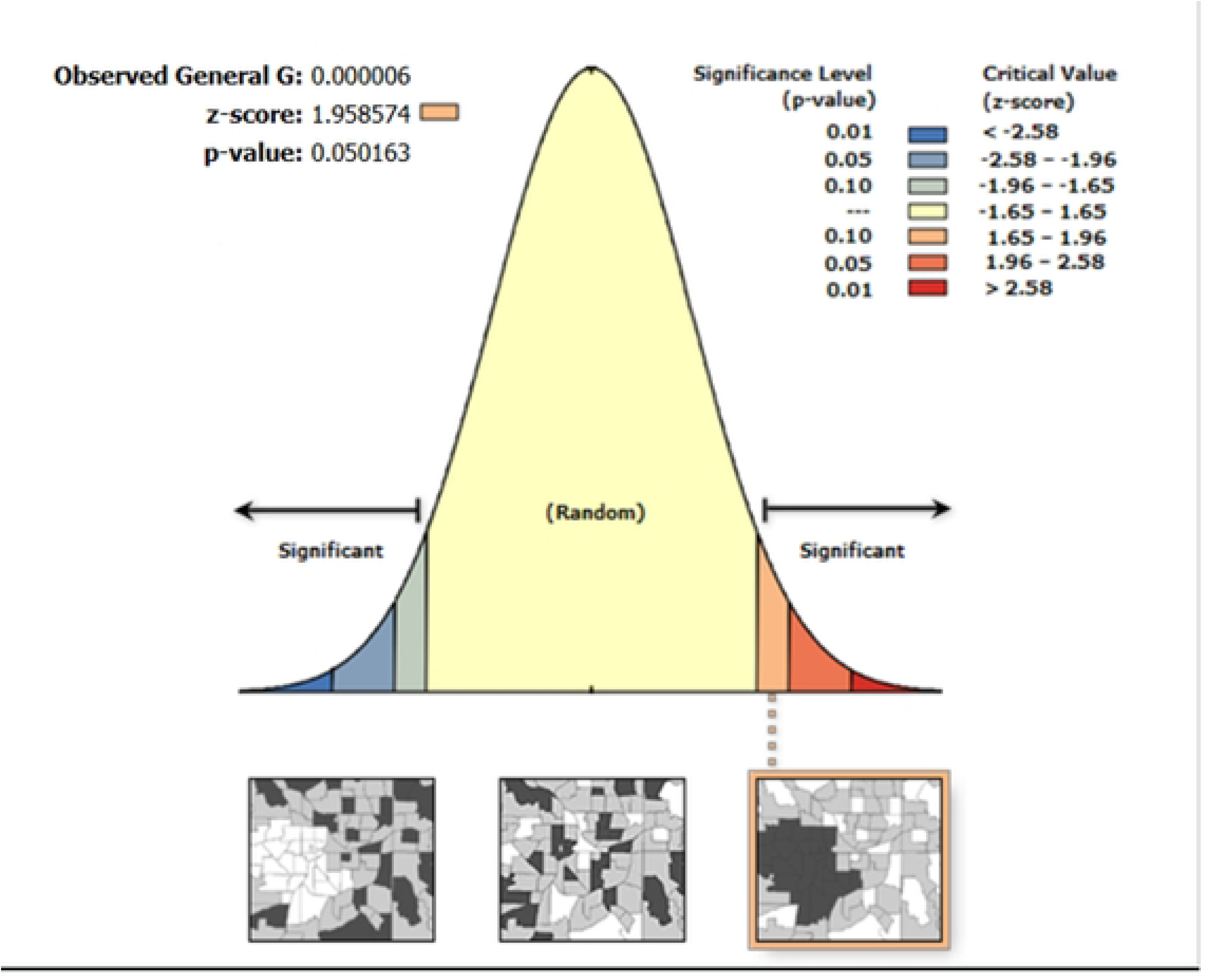
Global Spatial Autocorrelation (Moran’s I) of Diarrhea Prevalence Among Children Under Five in Mozambique, DHS 2022/2023.

#### High/Low hotspot analysis

The hotspot analysis using the Getis-Ord General G statistic revealed a Z-score of 1.95, a p-value of 0.05, and an observed General G value of 0.000. These results suggest that there is moderate but statistically significant clustering of high values across the study area. Since the p-value is exactly 0.05, this indicates that the spatial clustering is just statistically significant at the 5% level, and the positive Z-score points to the presence of hotspots—areas where high values of the studied variable are more concentrated than would be expected by chance. Although the observed General G value is close to zero, the Z-score and p-value together imply that there is a tendency for high values to be spatially clustered. Further local spatial analysis, such as the Getis-Ord Gi* statistic, is recommended to pinpoint specific hotspot locations within the country (Fig 2).

**Figure 2:**
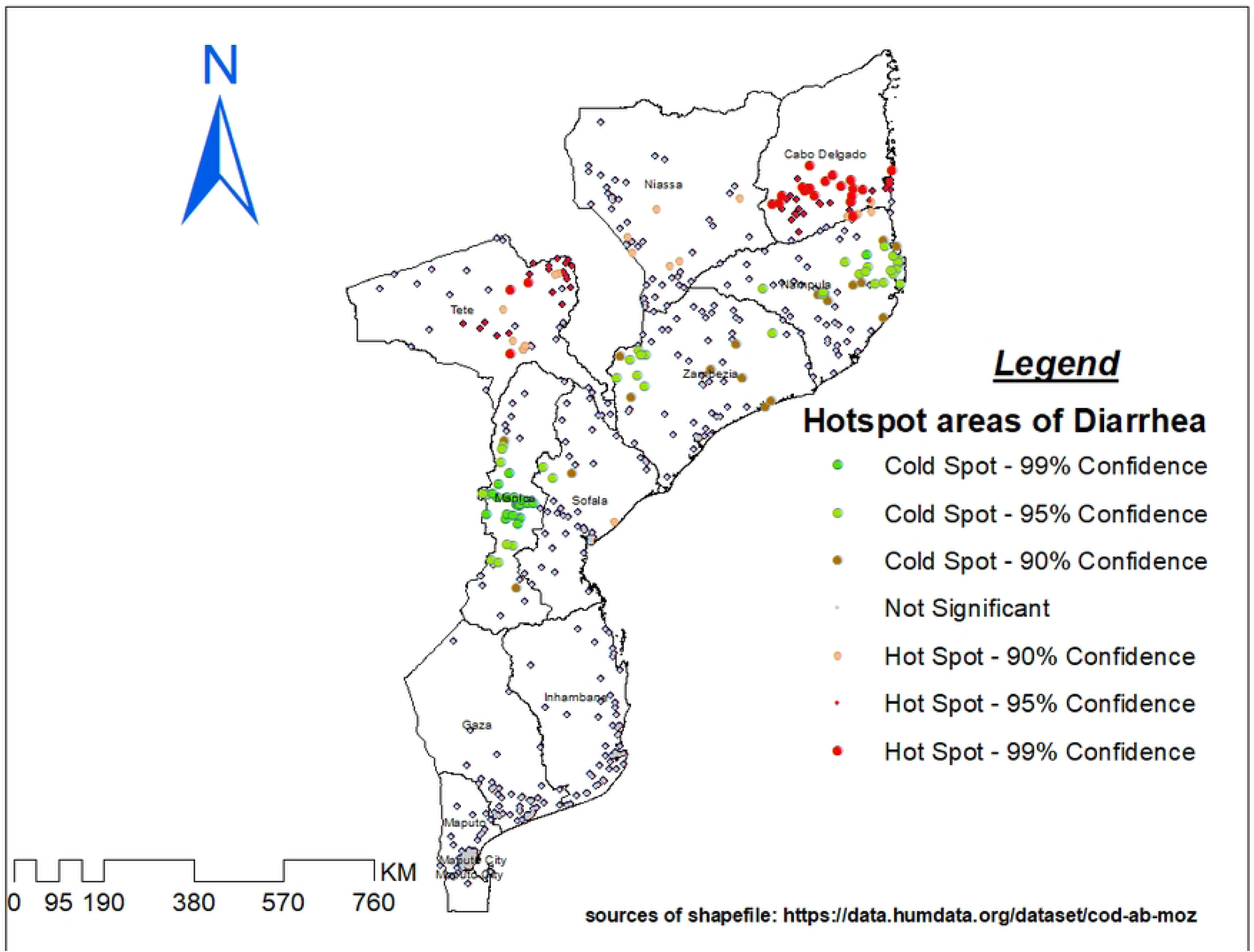
Global Spatial Autocorrelation (Moran’s I) of Diarrhea Prevalence Among Children Under Five in Mozambique, DHS 2022/2023.

#### Hotspot analysis

Hot spot analysis was performed to identify high-risk areas of under-five diarrhea in Mozambique. The red color (hot spot) indicates significant risky areas and is found in Northern Region (Cabo Delgado, Niassa, Nampula, Zambezia) and Central Region (Tete, Manica, Sofala), whereas the blue color indicates less risky areas (clod spot) of under-five diarrhea and is observed in Southern Region (Gaza, Inhambane, Maputo Province, Maputo City) and the Coastal Region (Fig 3).

**Figure 3:**
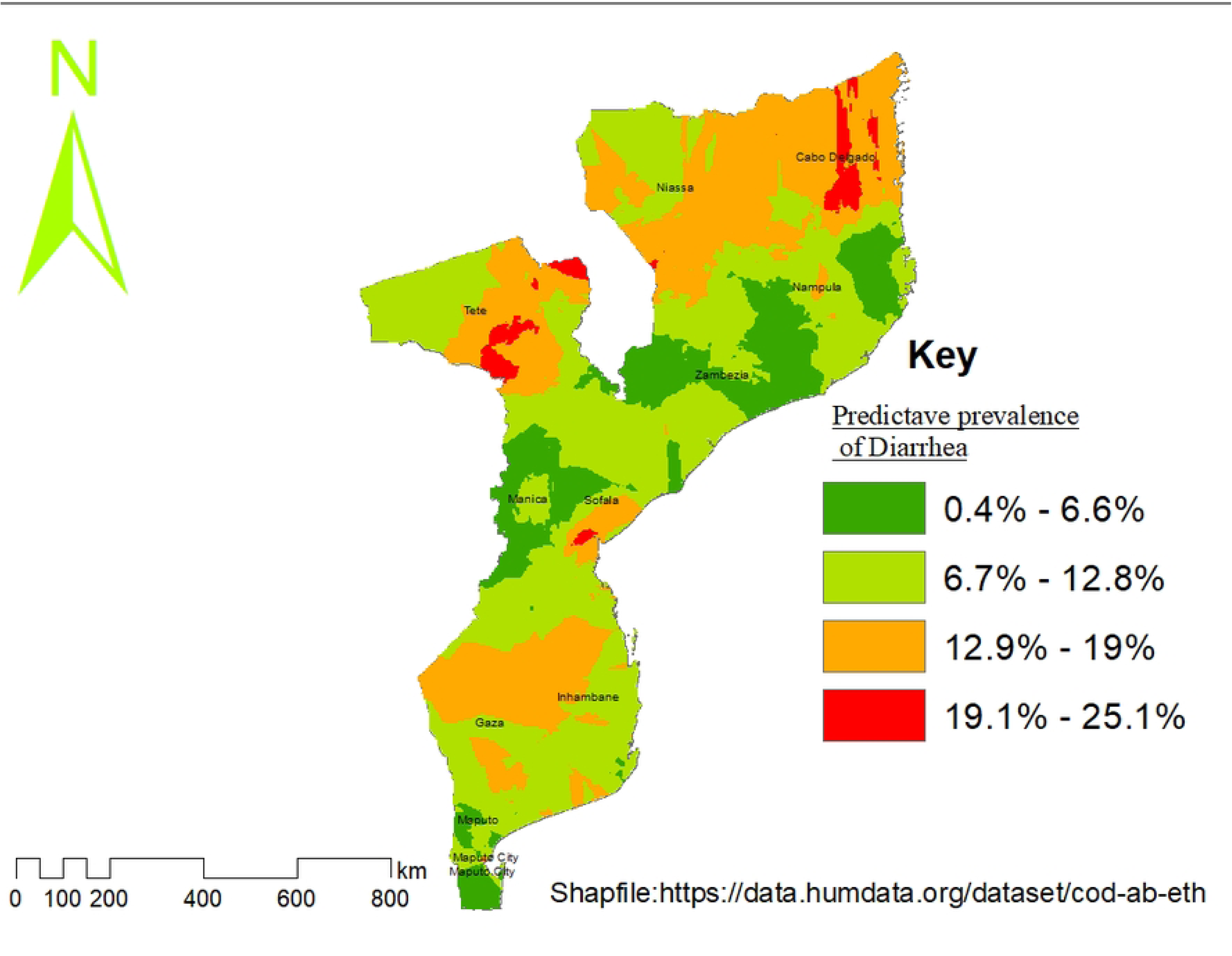
High and Low Hotspot Analysis of Diarrhea Prevalence Among Under-Five Children.

#### Ordinary Kriging Interpolation Map

The spatial interpolation results from the 2022/2023 Mozambique Demographic and Health Survey (DHS) revealed that the highest predicted prevalence of diarrhea illness is located in the northern regions (Niassa and Cabo Delgado). Maputo City, found in the south, also appears as an area with high predicted prevalence. Moderate prevalence levels are observed in the central regions (Tete, Gaza, Inhambane, and Sofala). The lowest predicted prevalence of diarrhea among under five children are detected in southern regions, especially in Maputo Province, Nampula, Manica, and Zambezia (Fig 4).

**Figure 4:**
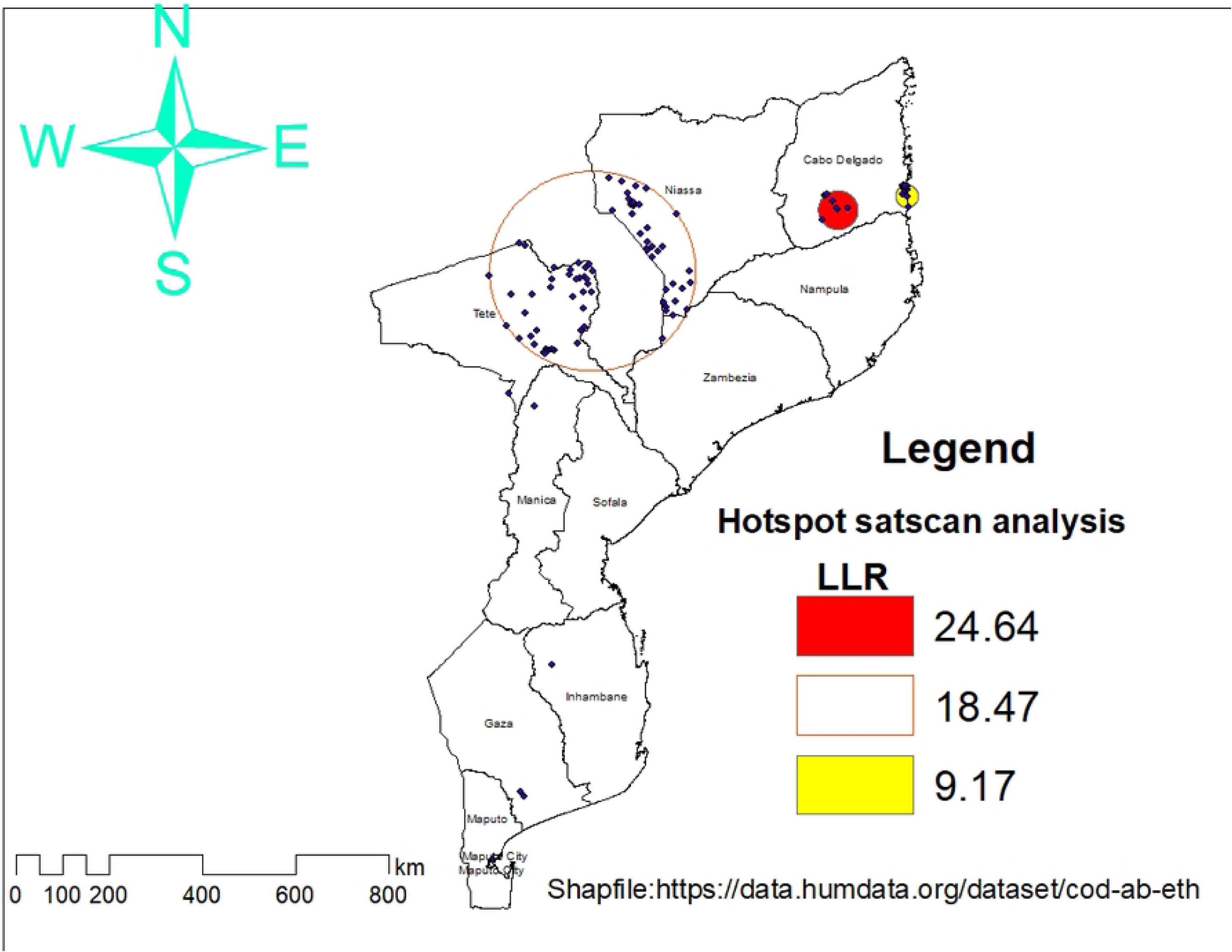
Spatial Interpolation of Diarrhea Prevalence Among Children Under Five in Mozambique Using Ordinary Kriging, DHS 2022/2023.

### SaTScan Cluster Detection (Spatial Scan Statistics)

According to the SaTscan analysis, under-five diarrhea cases in Mozambique are not randomly distributed, but are concentrated in specific high-risk areas across 93 geographic areas (within two clusters), primarily in the north (Niassa and Cabo Delgado) and parts of the central and southern regions (Tete and Gaza). The first cluster includes 10 locations and is centered around coordinates (13.42°S, 39.22°E) with a radius of 41.38 km. This cluster has a relative risk of 2.79, indicating that children in this area are nearly three times more likely to experience diarrhea compared to those outside the cluster. The log likelihood ratio is 24.65, and the p-value < 0.01, confirming a highly significant clustering. The second cluster includes 83 locations and is centered around coordinates (14.59°S, 34.49°E) with a radius of 213.63 km. This cluster has a relative risk of 1.61. he logs likelihood ratio is 18.47, with a p-value <0.01, also indicating strong statistical significance. From three up to seven spatial clusters like (Maputo Province, Zambezia, Manica, and Nampula) were located outside statistically significant clusters. These areas reflect a lower predicted prevalence of diarrhea among children under five (Fig 5, Table 4).

**Figure 5:**
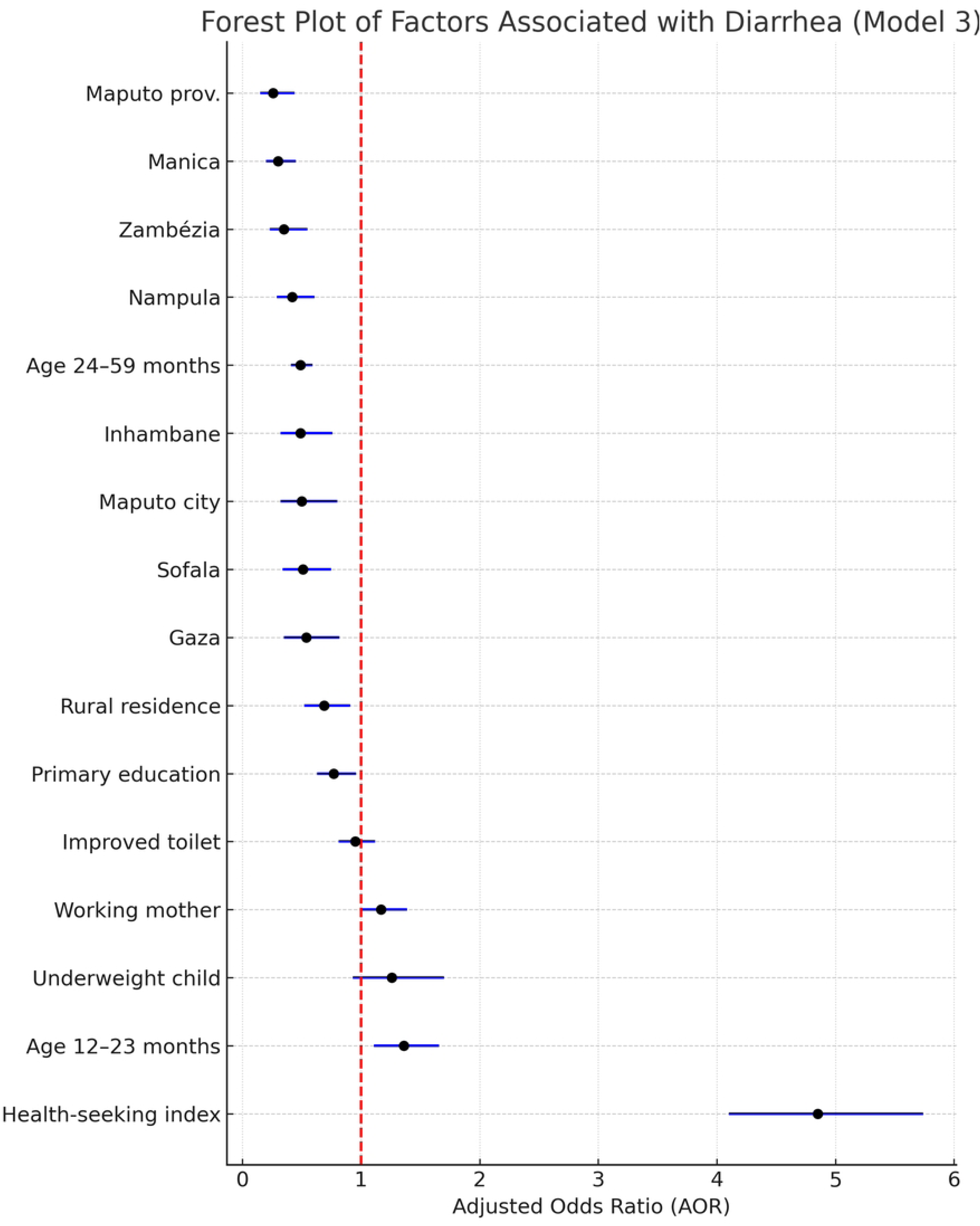
Significant Spatial Clusters of Childhood Diarrhea Identified by SaTScan in Mozambique, DHS 2022/2023.

**Table 4:** Statistically Significant Spatial Clusters of Diarrhea Among Children Under Five Identified by SaTScan in Mozambique, DHS 2022/2023.

## Discussion

Nationally, the prevalence of diarrhea among under-five children in Mozambique was 8.8% (95% CI: 7.8–9.6), based on the 2022/2023 DHS data, which shows a slight decline from previous surveys 10.5% in 2011 and 14.2% in 2003(8). Diarrhea remains a leading cause of under-five mortality in low- and middle-income countries, particularly when prevalence exceeds 5% without effective intervention (37). Mozambique’s 8.8% prevalence indicates a moderate public health burden, although it is lower than some sub-Saharan countries(27, 38). This underscores the need for multisectoral strategies to reduce incidence and improve treatment. As part of SDG 3, WHO and UNICEF promote integrated interventions; breastfeeding, rotavirus vaccination, zinc, ORT, and WASH(38, 39). While Mozambique shows progress, targeted efforts are still needed in high-burden districts.

The 2022/2023 DHS analysis shows that **regional disparities** in under-five diarrhea prevalence across Mozambique, consistent patterns seen in Uganda and Malawi, where rural-urban inequality, poverty, and low education contribute to high rates (40). Niassa (14.6%) and Cabo Delgado (14.1%) had the highest prevalence. It might be linked to poor health infrastructure, insecurity, and inadequate sanitation challenges worsened by ongoing conflict in northern regions (41, 42). Surprisingly, Maputo City also reported a high rate (13.4%), likely due to overcrowded informal settlements with poor sanitation and limited piped water access (43). As in other urban areas across sub-Saharan Africa, urban poverty remains a key driver of diarrheal disease through overcrowding and unsafe hygiene conditions(44).

**In contrast, Maputo Province recorded the lowest prevalence** (5.2%), The province may have better infrastructure and health service coverage compared to other provinces. The region’s high affluence and substantial health system investment could potentially enhance child health outcomes(45). Similarly, the low diarrhea prevalence in Zambézia (5.5%), Manica (5.9%), and Nampula (6.5%) is unexpected given their historically high burden. This may reflect recent gains in WASH efforts, expanded community outreach, or seasonal factors that temporarily lowered transmission during the survey period(46).

Provinces with **moderate prevalence,** such as Tete (12.3%), Gaza (12.0%), Inhambane (11.1%), and Sofala (10.4%), present a mixed picture. The regions in question are subjected to varying degrees of climatic stress, such as floods and droughts, which significantly impact their water safety and hygiene practices (47). The study reveals persistent environmental and behavioral risk factors in childhood diarrhea in Mozambique, highlighting the need for tailored interventions in northern regions (lNiassa and Cabo Delgad) and lower prevalence provinces (42).

The **prevalence of diarrhea** among under-five children in urban areas is 11.7% (95% CI: 9.97%–13.36%), while it is 7.5% in rural areas (95% CI: 6.56%–8.66%). Urban children in Mozambique face higher diarrhea risk than rural peers, likely due to poor sanitation, unsafe water, and inadequate waste management in informal settlements(27). These conditions increase environmental contamination and diarrheal pathogen transmission. Overcrowding in urban slums further amplifies fecal-oral spread due to close contact and poor hygiene facilities(48). Urban areas face increased diarrhea risk due to public sanitation issues, food contamination, and poor storage, with urban caregivers reporting more cases, especially in sub-Saharan Africa(49, 50). The analysis highlights Mozambique’s urban vulnerability, particularly among under-five children, and calls for urgent urban-targeted WASH interventions like safe water access, improved sanitation, and health education. UNICEF’s 2019 analysis reveals urban slums often have worse sanitation than rural villages, despite rural children’s 31% lower diarrhea prevalence(51). Certain regions, including Maputo Province, Manica, and Zambézia, may lower diarrhea rates in children due to improved health programs and WASH coverage, supported by international NGOs(27). The current findings in Mozambique align with regional patterns across Africa, showing diarrhea is more prevalent in conflict-affected, poor, and infrastructure-deficient areas, both rural and urban(50). The SaTScan analysis revealed considerable clusters of high-risk locations for diarrhea. The most likely cluster (Cluster 1), with a relative risk (RR) of 2.79, contains the northern districts of Niassa and Cabo Delgado. The second most likely cluster (Cluster 2), with an RR of 1.61, covers central Mozambique. The study identifies clusters of high diarrhea prevalence in regions with limited access to clean water and sanitation, supporting targeted intervention in these areas, similar to Ethiopia and Nigeria(4, 52).

Children aged 12–23 months had significantly higher odds of experiencing diarrhea (AOR = 1.36; 95% CI: 1.11–1.66) compared to those under one year, while children aged 24–59 months were significantly less likely to have diarrhea (AOR = 0.49; 95% CI: 0.41–0.59). Previous studies, have identified a vulnerability linked to weaning practices, increased mobility, and exposure(53-55). Similar trends were reported in studies from Ethiopia and Nigeria(56, 57), and a pooled analysis across SSA found diarrhea incidence peaking between 12 and 23 months(58). This study is in line with studies conducted at Tanzania and Ethiopia showed a lower likelihood of developing diarrhea, possibly due to immunity from earlier exposures(54, 55, 59, 60).

Children of employed mothers have 1.2 times higher diarrhea odds due to reduced childcare time and reliance on alternate caregivers, with maternal employment also linked to increased risk of childhood illness(61). This may be due to reduced time for child care, feeding, hygiene, and health-seeking practices when mothers are engaged in outside work(62-64). Evidence from Nigeria and Ethiopia supports reduced breastfeeding and increased environmental risks ((65, 66), possibly due to time constraints in low-income settings. Children of mothers with primary education had significantly lower odds of diarrhea (AOR = 0.77; 95% CI: 0.63–0.96) compared to those whose mothers had secondary education or higher. Basic educational attainment significantly improves child health outcomes, including nutrition, healthcare utilization, hygiene practices, and child diarrhea, despite a seemingly small odds reduction in East Africa like Mozambique and Ghana (67, 68). Higher health-seeking behavior index is linked to nearly 5 times higher diarrhea rates, possibly due to mothers actively seeking healthcare. A similar phenomenon was observed in Kenya(69). This may reflect reporting bias rather than actual disease burden.

The study found that children in Maputo Province had a 74% lower odds of diarrhea, possibly due to improved infrastructure and health services, consistent with study in southern Mozambique(70). Maputo City demonstrated a decrease in odds (AOR = 0.50), indicating improved WASH access and vaccine coverage, as reported by UNICEF (27). Zambézia and Manica showed significantly lower odds (AOR = 0.35 and 0.30) of developing a disease, possibly due to the effectiveness of hygiene campaigns and maternal health programs(27). Inhambane and Gaza experienced reduced odds (AOR = 0.49 and 0.54) of having improved toilets, similar to central-southern Mozambique where access to improved toilets was higher(70).

Children living in several regions of Mozambique had significantly lower chances of having diarrhea compared to those in Niassa. Specifically, the risk was lower in Nampula (58% lower, AOR = 0.42), Zambézia (65% lower, AOR = 0.35), Manica (70% lower, AOR = 0.30), Sofala (49% lower, AOR = 0.51), Inhambane (51% lower, AOR = 0.49), Gaza (46% lower, AOR = 0.54), Maputo Province (74% lower, AOR = 0.26), and Maputo City (50% lower, AOR = 0.5. The study indicates disparities in environmental and healthcare conditions across regions, possibly influenced by factors such as water safety, infrastructure, climate, and localized interventions. Studies from Kenya, Tanzania, and other parts of Mozambique revealed comparable spatial disparities (6, 8). The study found that significant variation in diarrhea risk was due to cluster-level differences, with an ICC decline from 15.2% to 8.5%, MOR decreasing from 2.08 to 1.69, and PCV reaching 49.2%. The model’s variables were found to explain nearly half of the variation between clusters. Similar ICC and MOR estimates were found in multilevel studies conducted in Ethiopia and Nigeria (71, 72) (Table 5).

**Table 5:**
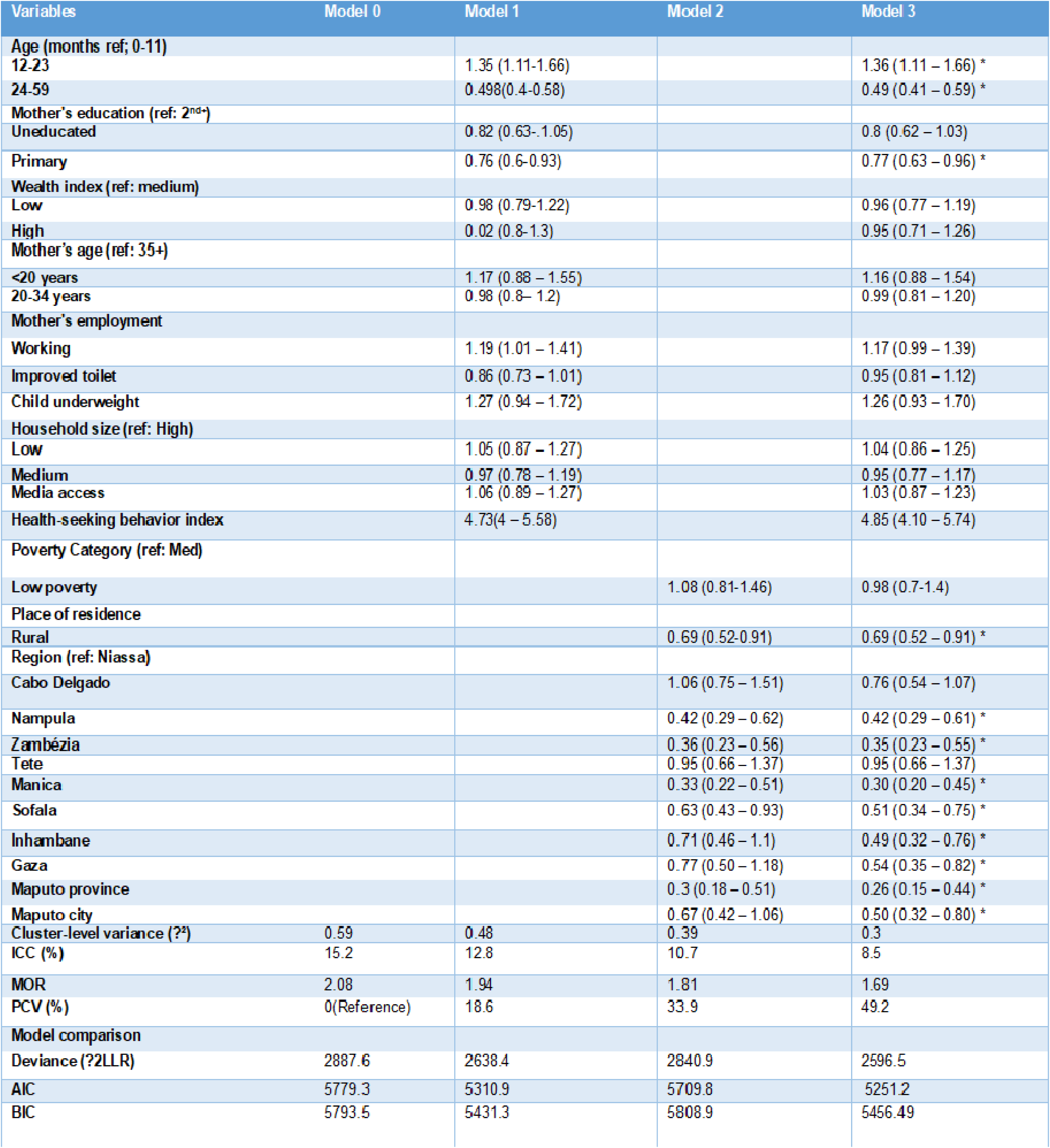
Multilevel Logistic Regression Models for Factors Associated with Diarrhea Among Children Under Five in Mozambique, DHS 2022/2023.

The 2022/2023 Mozambique DHS data offers valuable insights that uses a nationally representative design, standardized questionnaires, and geospatial and biomarker data to assess diarrhea risk in Mozambique. It provides robust statistical power, precise estimates, and subgroup analyses. The study’s recency and integration of contextual factors make it a strong foundation for evidence-based interventions. However, its cross-sectional design, reliance on maternal recall, GPS displacement, lack of pathogen-specific data, seasonal bias, unmeasured factors, and spatial aggregation may cause ecological fallacies.

## Conclusions

Diarrhea continues to pose a moderate public health challenge for under-five children in Mozambique, with clear disparities across regions. Factors like child age, maternal education, rural vs urban residence, and geographic location influence the risk of childhood diarrhea. Future interventions should focus on age and area-specific risk factors. Targeted interventions include expanding maternal education, improving WASH infrastructure, and enhancing maternal support systems, particularly in high-burden regions such as Niassa and Cabo Delgado.

## Data Availability

No

https://dhsprogram.com

## Consent for publication

Not applicable

## Availability of data and materials

The datasets and materials used in this study are publicly available from the Demographic and Health Surveys (DHS) Program website at www.dhsprogram.com, specifically for the Mozambique 2022/2023 survey. Additional data or materials generated or analyzed during this study can be obtained from the corresponding author upon reasonable request.

## Authors Contribution

Conceptualization, Formal Analysis, Methodology, Investigation=Thomas Kidanemariam Yewodiaw, Mihret Getnet

Resources, Software Validation,Visualization: Thomas Kidanemariam, Mequanent Dessie Bitewa,, Hiwot Tezera Endale

Writing – Original Draft: Thomas Kidanemariam, Hiwot Tezera Endale, Mihret Getnet

Writing – Review & Editing: Thomas Kidanemariam, Mihret Getnet,

## Ethics approval and consent to participate

The data used in this study are from the Demographic and Health Surveys (DHS) Program. Although public access to the DHS datasets is currently paused, we obtained the required data during a prior approved request as part of a previous research project. The datasets are available from https://dhsprogram.com upon reasonable request and approval from the DHS Program.

## Abbreviations

U5C; Under-Five Children SaTScan; Spatial and Space-Time Scan Statistics, GPS; Global Positioning System, WHO; World Health Organization, SSA; Sub-Saharan Africa, UNICEF; United Nations Children’s Fund, ICC; intraclass correlation coefficient, MOR; median odds ratio, PCV; proportional change in variance, MDHS, Mozambique Demographic Health Surveys; KR; Kids Recode (DHS child-level dataset), EAs, enumeration areas; AOR; Adjusted Odds Ratio, CI; Confidence Interval, p-value; Probability value (significance level).

## Acknowledgments

We would like to thank the Mozambique National Institute of Statistics (INE) for providing access to the dataset used in this study, and the DHS Program for granting permission to download the necessary supporting materials.

